# Genome-wide analysis of screen behaviors among adolescents identifies novel loci and overlap with educational attainment and mental disorders

**DOI:** 10.1101/2025.01.07.25320110

**Authors:** Evgeniia Frei, Tahir Tekin Filiz, Oleksandr Frei, Robert Loughnan, Piotr Jaholkowski, Nora R. Bakken, Viktoria Birkenæs, Alexey A. Shadrin, Helga Ask, Ole A. Andreassen, Olav B. Smeland

**Affiliations:** Centre for Precision Psychiatry, Institute of Clinical Medicine, University of Oslo, Oslo, Norway; Division of Mental Health and Addiction, Oslo University Hospital, Oslo, Norway; Centre for Bioinformatics, Department of Informatics, University of Oslo, Oslo, Norway; Center for Population Neuroscience and Genetics, Laureate Institute for Brain Research, Tulsa, Oklahoma, USA; Center for Human Development, University of California, La Jolla, California, USA; PsychGen Centre for Genetic Epidemiology and Mental Health, Norwegian Institute of Public Health, Oslo, Norway; PROMENTA Research Center, University of Oslo, Oslo, Norway

## Abstract

Technological devices play a central role in adolescents’ life. Despite concerns about negative effects of excessive screen time on mental health and development, there is little knowledge of fundamental features of screen behaviours and underlying genetic architecture. Using self-reports from adolescents (14-16 years old) in the Norwegian Mother, Father, and Child Cohort Study (MoBa, *n* = 18 490), we performed genome-wide association analysis for four screen behaviors: time spent 1) watching movies/series/TV; 2) gaming; 3) sitting/lying down with PC, mobile or tablet; and 4) communicating with friends on social media. The resulting summary statistics were analysed using the conditional false discovery rate (condFDR) approach to increase genetic discovery. We also estimated SNP-based heritabilities of the screen behaviours and the genetic correlations with six major psychiatric disorders (schizophrenia, bipolar disorder, major depressive disorder, autism spectrum disorder, attention-deficit hyperactivity disorder, alcohol use disorder), and educational attainment. The screen-based phenotypes displayed significant SNP-based heritabilities (0.048–0.12). We also observed significant genetic correlations between screen behaviours and psychiatric disorders (r_g_ range: 0.21–0.42). Educational attainment demonstrated significant negative genetic correlation with screen behaviours, most strongly with social media use (r_g_ = −0.69). CondFDR analysis identified three novel loci associated with social media use. Thus, we show that screen behaviors are heritable, polygenic traits that partly share genetic signal with mental disorders and educational attainment. Future studies and larger samples are required to clarify causal relationships between these traits and disorders, and to validate the identified genetic loci associated with social media use.

## Introduction

Technological devices have become an integral part of adolescents’ life. The majority of young people spend several hours per day on screen-based activities, and reports indicate that the numbers continue to increase (1,2). Child and adolescent use of technology outpaces our understanding of the fundamental features and health impact of screen behaviours, and additional research is needed to guide children’s access to social media, as well as propose the optimal use of screen devices in primary and secondary education. In the 1990s, the first evidence of genetic influence on television watching emerged, challenging the notion that it was a pure “environmental factor” (3). Extensive research has since confirmed that screen behaviours are heritable traits (4), including substantial twin heritability estimates of gaming behaviour (19%-63%) (5), compulsive internet use (48%) (6), and problematic internet use (58%-66%) (7) among adolescents. Recent work investigated SNP-based heritability (*h^2^_SNP_*) of screen behaviours (TV/video watching, gaming, total screen time) in the Adolescent Brain Cognitive Development Study, and reported estimates that varied from zero to 10-18%, depending on the screen subtype (8).

Despite this, the genetic architecture of screen behaviours among adolescents is poorly understood with a lack of studies detecting specific single nucleotide polymorphisms (SNPs) (9). A recent genome-wide association study (GWAS) revealed several SNPs significantly associated with internet addiction disorder in adults (10). Further, GWASs based on middle-aged individuals in the UK Biobank (UKB) cohort identified SNPs associated with television watching and leisure computer use (11). However, it is unclear whether these results can be generalized to adolescents, who spend more time on screen devices than any other age group (12).

Parallel to the widespread use of digital devices among young people, youth mental health problems are on the rise (13–15). While excessive use of screen devices has been linked to negative mental health outcomes, the proposed explanations vary greatly (16–19). Psychiatric disorders in children and adolescents are affected by genetic factors (20–22), and the potential of shared genetic determinants underlying screen time and mental health problems in children and adolescents is a novel, emerging research topic (23). Recent studies have indicated that genetic confounding may account for a substantial part of the phenotypic association between screen time and mental health (24,25), and that major psychiatric disorders and screen behaviours may share a common genetic basis (23).

Furthermore, a growing body of evidence suggests that increased screen time could affect academic performance in children and adolescents (26). For example, television watching, gaming, and social media use are associated with worse academic performance (27–29). Evidence also suggests that prolonged screen time could contribute to a diminished capacity for sustained attention and a heightened susceptibility to distractions (30). Finally, both twin studies and large genome-wide association studies have demonstrated that genetic factors are important for educational attainment (31,32), although the extent to which academic performance and screen behaviours share genetic underpinnings remains unclear.

In this study, we leveraged data from the Norwegian Mother, Father, and Child Cohort Study (MoBa) (33), a prospective population-based pregnancy cohort, to investigate the genetic architecture of screen behaviours among adolescents and their associations with key mental health traits and disorders. We aimed to identify specific genomic loci associated with screen behaviours in adolescents. To achieve this, we performed GWASs of four single screen behaviours (television watching, gaming, total screen time use, and social media use). We undertook extensive post-GWAS analyses, including estimating genetic correlations across screen behaviours and with six major psychiatric disorders (schizophrenia [SCZ], bipolar disorder [BP], major depressive disorder [MDD], autism spectrum disorder [ASD], attention-deficit hyperactivity disorder [ADHD], and alcohol use disorder [AUD]), as well as educational attainment (EA). In addition, we analysed the resulting summary statistics using the conditional false discovery rate (condFDR) approach to improve statistical power and genetic discovery (34,35).

## Methods

### Study sample

MoBa is a population-based pregnancy cohort study conducted by the Norwegian Institute of Public Health (NIPH) (33). Participants were recruited from all over Norway from 1999-2008, and the women consented to participation in 41% of the pregnancies. The cohort includes approximately 114 500 children, 95 200 mothers and 75 200 fathers. For genotyping, blood samples for the children were taken from the umbilical cord after their birth. Biological samples were sent to NIPH where DNA was extracted by standard methods and stored. Genotyping of the MoBa cohort was conducted through multiple research projects spanning several years, involving various selection criteria, and genotyping centres. Full details about the genotyping and quality control procedures are provided elsewhere (36).

The current study is based on version 12 of the quality-assured data files released for research in January 2019, including all adolescents (14-16 years of age) with relevant phenotypic and genetic data available (*n* = 18 490). Questionnaire data were collected between 2017-2023. Data of participants who withdrew their consent before August 2024 were not included in the analyses. The establishment of MoBa and initial data collection was based on a license from the Norwegian Data Protection Agency and approval from The Regional Committees for Medical and Health Research Ethics (REK). The MoBa cohort is currently regulated by the Norwegian Health Registry Act, and the current study was approved by REK (2016/1226/REK sør-øst C).

### Screen behaviors

We used single item self-reports about activities during leisure time from the MoBa Q-14year questionnaire. Specifically, adolescents reported how much time they spent on the following screen-based activities: 1) watching movies/series/TV; 2) playing games on PC, TV, tablet, mobile, etc.; 3) sitting/lying down with PC, mobile, or tablet (irrespective of activity); 4) communicating with friends on social media). For each activity, responses were coded from 1 (never/rarely) to 6 (7 hours or more). Detailed information about study variables is presented in the Supplementary Table 1.

### Genome-Wide Association Analyses

GWASs were conducted using an additive multivariate linear regression model with PLINK2 (37) (v2.00a5.10LM). To ensure that screen use patterns reflect the general adolescent population as accurately as possible, the sample was restricted to participants without a high likelihood of severe disability. In line with another adolescent cohort study (38), we therefore excluded individuals with current diagnosis of SCZ (F20-F29), mental impairment/intellectual disability (F70-F79), alcohol/substance use disorder (F10-F19), and ASD (F84), based on the data from the Norwegian Patient Registry (NPR), which contains diagnoses registered in specialist healthcare services. Moreover, subjects with ambiguous sex and subjects with chromosomal abnormalities were removed from analyses. This resulted in a sample of 17 945 individuals. For each pair of participants with a kinship coefficient greater than 0.05, one member was randomly excluded from analyses, resulting in a sample of 16 027 unrelated individuals. The first twenty genetic principal components (PCs), age, sex, and genotyping batch (N = 26, as factors) were used as covariates. The analyses were restricted to individuals of European ancestry selected as described previously (36). All summary statistics underwent quality control and were cleaned using the cleansumstats pipeline (39). Analyses were conducted using singularity containers (40).

### Sensitivity analysis

To ensure that the presence of psychiatric diagnoses in the study sample does not confound the results, we performed a sensitivity analysis and re-estimated all genetic correlations using GWAS on screen phenotypes based on subsample of participants without a history of any psychiatric disorder, according to the NPR data. Of the 18 490 participants with relevant phenotypic and genetic data available, 3705 (20.04%) had at least one registered psychiatric diagnosis (see Supplementary Table 2 for more detailed information).

### Conditional False Discovery Rate (condFDR) analyses

To improve statistical power and genetic discovery, we used the condFDR approach. Briefly, it boosts GWAS discovery by leveraging overlapping SNP associations between two GWASs to re-rank the test statistics in a primary phenotype conditional on the associations in a secondary phenotype (34,35). In our study, the primary phenotypes were the four screen time measures, with educational attainment as a secondary phenotype (32). To facilitate evaluation of identified loci in the UK Biobank, we excluded this cohort from the EA summary statistics. To visualize cross-trait enrichment we used conditional QQ plots, which show *p*-value distributions for a primary trait for all SNPs, and for SNP strata set by their association with a secondary trait. For QQ plots production, we excluded variants within regions with complex linkage disequilibrium (LD) structure (MHC region: chr6:25119106–33854733, and 8p23 inversion: chr8:7200000–12500000, GRCh37 coordinates). Successive leftward deflection of the variant strata with increasing significance in the conditional phenotype in both directions suggests strong cross-trait enrichment. The FDR significance cut-off for condFDR was set at 0.01, in line with the previous literature (34,35).

### Evaluation of the identified Loci in an Independent Sample

We used GWAS results from the UKB cohort on leisure television watching (TV-UKB) and leisure computer use (PC-UKB) to test whether our results can be supported by data from an independent sample (11). For this purpose, we checked whether effects of the lead SNPs identified by condFDR analysis are consistent between the MoBa and UKB data sets. Additionally, we obtained the *p*-values of the lead SNPs from the MoBa cohort in the UKB sample.

### Functional Analyses

Genomic loci were defined by identifying independent significant SNPs with condFDR < 0.01 that were not in close LD with each other (r^2^ < 0.60), according to the FUMA protocol(41). Lead SNPs were then defined by the independent significant SNPs with r^2^ < 0.1 in approximate LD. Candidate SNPs were defined as all SNPs with condFDR < 0.10 and in LD (r^2^ ≥ 0.60) with an independent significant SNP. The loci borders were set by identifying all candidate SNPs in LD (r^2^ ≥ 0.6) with one of the independent significant SNPs in the locus. Loci < 250_kb apart were merged, and the lead SNP of the merged locus was selected as the SNP with the most significant condFDR value. Overlapping signals within complex LD regions were represented by one independent lead SNP only. All LD r^2^ values were obtained from the 1000 Genomes Project European-ancestry haplotype reference panel (42).

Using FUMA (41), all candidate SNPs with condFDR < 0.10 were functionally annotated with combined annotation-dependent depletion (CADD) scores to predict deleterious SNP effects on proteins, RegulomeDB scores to predict the likelihood of regulatory function of a SNP, and chromatin state scores to predict transcriptional effects. Candidate SNPs were aligned to genes using three different strategies implemented in FUMA (positional gene mapping, expression quantitative trait locus mapping, and chromatin interaction mapping). The lead SNPs were also mapped to putative causal genes using the Variant to Gene (V2G) tool from the open-source OpenTargets Genetics (43). The OpenTargets Genetics platform was also used to inspect associations of the mapped genes with other phenotypes. To estimate expression of the mapped genes in human brain, we used the open-source Brain RNA-Seq Database (44).

### Estimation of SNP-Based Heritabilities and Genetic Correlations

*h^2^_SNP_* of screen behaviors were estimated from the GWASs summary statistics using linkage disequilibrium score regression (LDSC) (45). To perform more precise estimation of *h^2^_SNP_* using individual genotype data, we conducted GCTA-GREML analysis (46,47).

We also applied bivariate LDSC (45) to estimate genetic correlations (r_g_) across screen behaviours and with six major psychiatric disorders (SCZ, BP, MDD, ADHD, ASD, AUD) (48–53), as well as EA (32). Genetic correlations were estimated in the main study sample, as well as in the subsample of participants without a history of a psychiatric disorder. We also estimated genetic correlations between the screen behaviors in MoBa and TV watching and leisure computer use in the UKB cohort.

We used the set of LD scores provided by the software’s creators, based on the 1000 Genomes Project’s European sample. Summary statistics from external GWASs of SCZ, BP, MDD, ADHD, ASD, AUD, and EA were harmonized using the cleansumstats pipeline. Additional SNP quality control routines were equivalent to the defaults employed with the LDSC *munge_sumstats.py* function. Following the recommended practices, we assumed no sample overlap.

Correlations are presented as the coefficient ± standard error. Original *p*-values are reported. Multiple testing correction was performed using the Benjamini-Hochberg procedure with FDR < 0.05. Analyses were done using Python ver. 3.9.5.

## Results

In total, 18 490 participants had relevant phenotypic and genetic data available, and 16 027 unrelated individuals were included in the genetic analysis. Basic demographic characteristics of the initial sample are presented in Table 1. Descriptive information on study variables is presented in the Supplementary Table 1.

**Table 1.** Basic demographic characteristics of the adolescent sample.

| **Demographic characteristics of the adolescent sample ($n = 18\,490$) from MoBa** |  |
| --- | --- |
| Age when questionnaire was answered, years, mean (SD) | 14.42 (0.51) |
| Sex assigned at birth, $n$ (%) | |
| Male | 8 621 (46.63%) |
| Female | 9 858 (53.32%) |
| NA | 11 (0.059%) |
| Gender identity, $n$ (%) | |
| Boy | 8 369 (45.26%) |
| Girl | 9 527 (51.53%) |
| Trans person | 42 (0.23%) |
| Do not know / Do not wish to answer | 254 (1.37%) |
| NA | 298 (1.61%) |

*NA indicate missing values.*

### GWASs of Screen Behaviours

The total number of missing values did not exceed 1.5% for any of the screen-based phenotypes. Detailed information about missing values, and sample sizes for each screen-based phenotype are available in the Supplementary Table 1.

There was no evidence of stratification artefacts or uncontrolled test statistic inflation in the results for any phenotype (Supplementary Figures 1-2). No SNP reached genome-wide significance in any of the four GWAS. A list of SNPs that reached the level of suggestive genome-wide significance (*p* < 1×10^−5^) for the four screen behaviors is given in the Supplementary Tables 4-7.

### SNP-Based Heritability

Analysis of the GWAS summary statistics indicated that the screen behaviours have significant nonzero *h^2^_SNP_* (Figure 1, Supplementary Table 3). LDSC results showed that *h^2^_SNP_* for television watching, gaming, and social media use was 0.066 (SE = 0.027, *p* = 0.0073), 0.070 (SE = 0.029, *p* = 0.0079), and 0.12 (SE = 0.031, *p* = 5.42×10^−5^), respectively. LDSC heritability estimate of the total screen time phenotype was not significantly different from zero.

**Figure 1.**
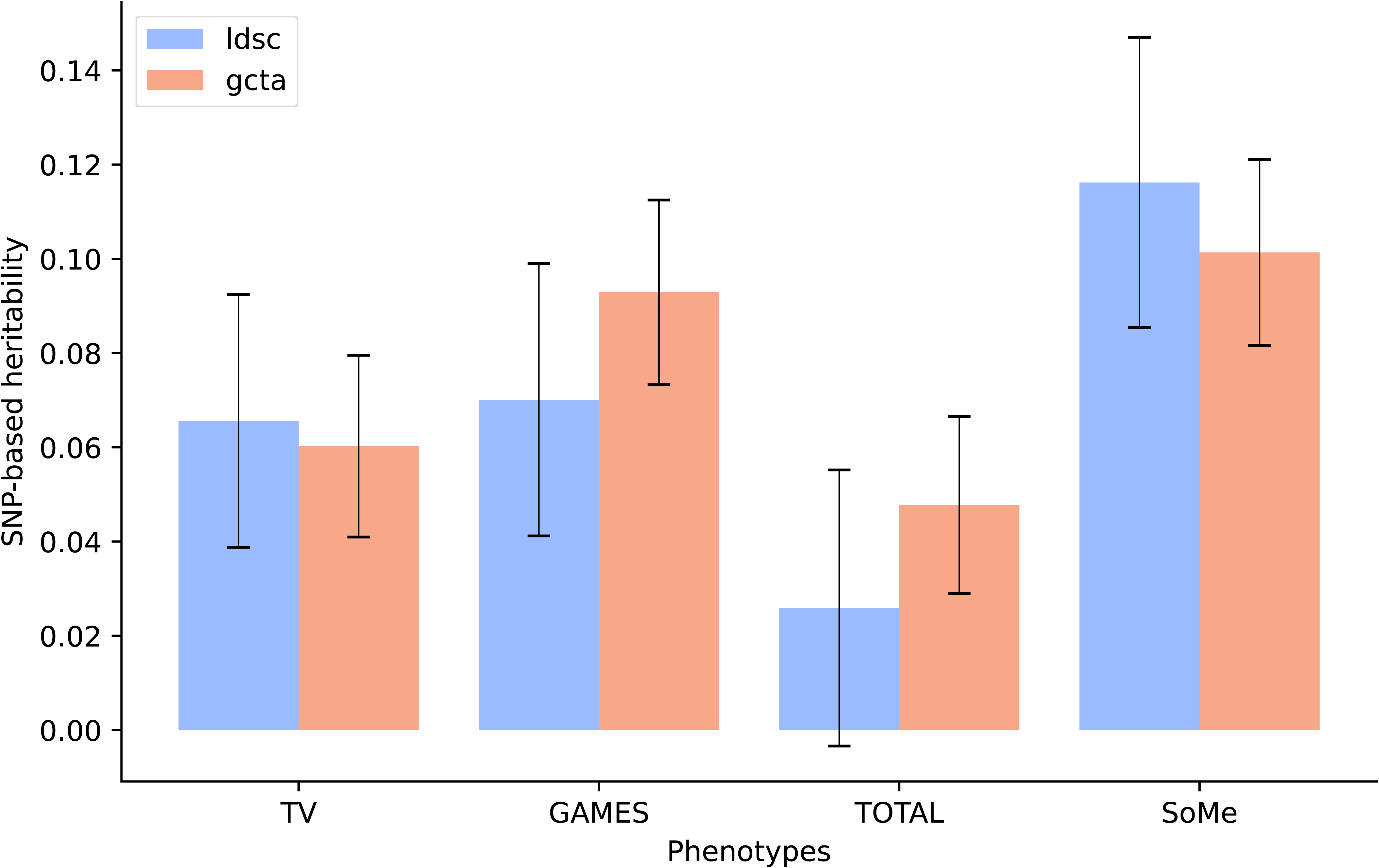
Single nucleotide polymorphism–based heritability (SNP-h^2^) estimates for screen behaviours, obtained with LDSC (blue bars) and GCTA-GREML (orange bars). Error bars indicate standard errors of the estimated values. TV: watching movies/series/TV; GAMES: playing games on PC, TV, tablet, mobile, etc.; TOTAL: sitting/lying down with PC, mobile or tablet; SoMe: communicating with friends on social media.

GCTA-GREML produced concordant heritability estimates. Specifically, *h^2^_SNP_* of television watching, gaming, and social media use was 0.060 (SE = 0.019, *p* = 0.0012), 0.093 (SE = 0.020, *p =* 4.55×10^−8^), and 0.10 (SE = 0.020, *p* = 2.01×10^−7^), respectively. Total screen time use *h^2^_SNP_* estimated with GCTA-GREML was significantly different from zero: 0.048 (SE = 0.019, *p* = 0.0048).

### Identification of Loci Associated with Social Media Use

Conditional QQ plots demonstrated enrichment of SNP-associations with social media use conditional on increasing levels of significance with EA (Supplementary Figure 3).

We leveraged this cross-trait enrichment using condFDR analyses and identified three LD-independent loci associated with social media use at condFDR < 0.01 (Figure 2). The lead SNPs in the identified loci were mapped to putative causal genes using the V2G tool from the Open Targets Genetics (43,54,55). The strongest signal was located at an intergenic variant (rs7110805, condFDR = 5.10×10^−5^), on chromosome 11 (Figure 3C). Its nearest gene is *MTMR2*, while the region also contains the genes *FAM76B* and *CEP57* (downstream). Three additional independent significant SNPs (rs1727149, rs10765775, rs1893057) were identified in this large region spanning more than 250 000 bp. The second strongest independent condFDR signal was an intergenic variant on chromosome 2 (rs359240, condFDR = 1.28×10^−3^, Figure 3A). No genes were residing in the direct vicinity of this SNP (the canonical TSS of the nearest gene *BCL11A* is located at 306 594 bp upstream), and only 34 SNPs were in strong LD (r^2^ > 0.6). Nevertheless, rs359240 has a high CADD score of 19.6, indicating deleteriousness(56). Finally, the condFDR analysis identified an ncRNC intronic variant on chromosome 4 (rs6848288, condFDR = 3.65×10^−3^, Figure 3B), with nearest protein-coding gene *SMARCAD1*. rs6848288 tags a broad region of associations, covering around 270 000 bp, and has 139 candidate SNPs in strong LD (r^2^ > 0.6). Besides *SMARCAD1*, this region also contains the *HPGDS* gene (upstream).

**Figure 2.**
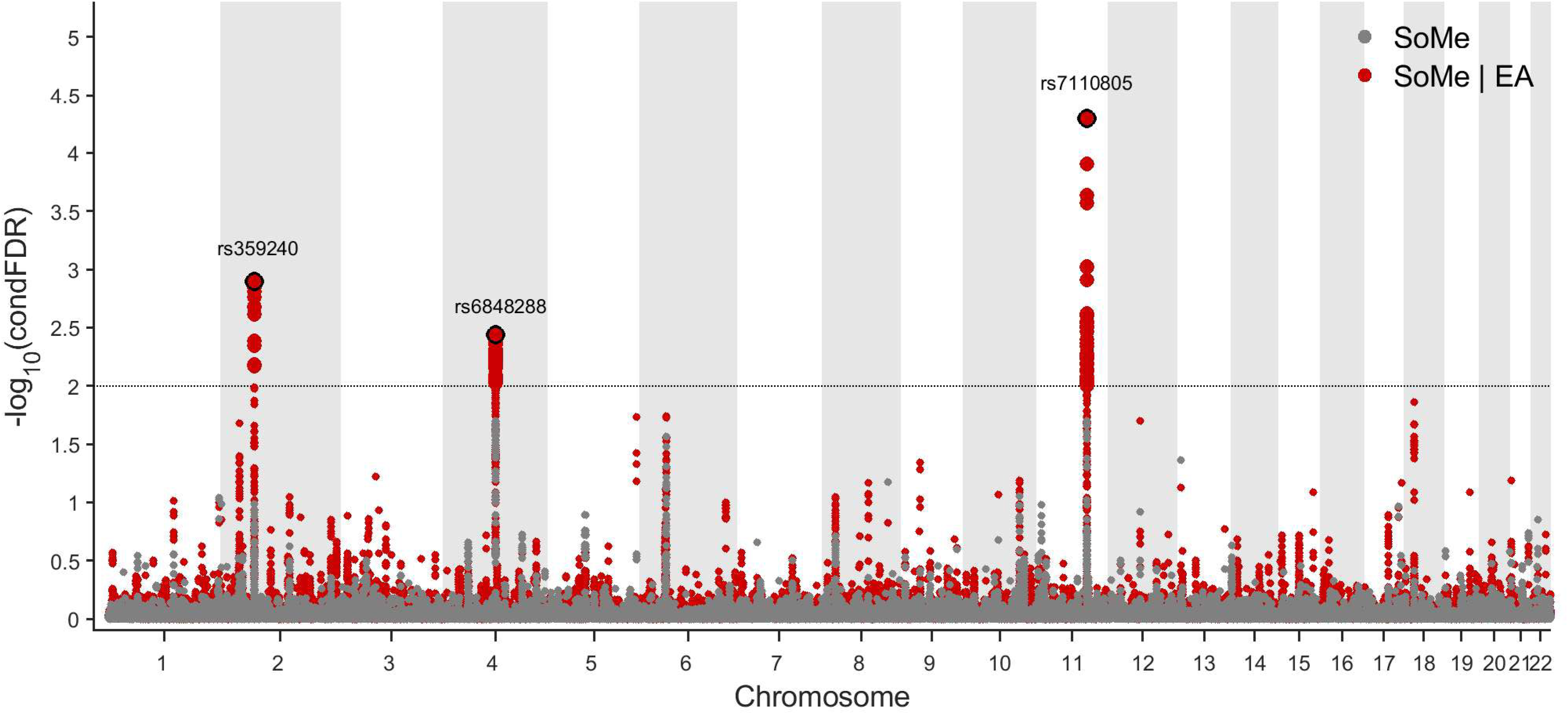
Common genetic variants significantly associated with social media use (SoMe) among adolescents in the MoBa sample. The variants were identified at conditional false discovery rate (condFDR) < 0.01 after conditioning on educational attainment (EA). The Manhattan plot displays the –log10 transformed condFDR values for each single-nucleotide polymorphism (SNP) on the y-axis with chromosomal positions along the *x*-axis. The small points represent non-significant SNPs, the bold points represent significant SNPs (condFDR < 0.01). Points corresponding to significant SNPs with lowest conditional FDR in each linkage disequilibrium (LD)-independent region (r^2^ > 0.10) have the rs-number written above it. The horizontal grey dotted line shows the significance threshold of condFDR (0.01). Gray dots stand for unconditional FDR values. SoMe: communicating with friends on social media.

**Figure 3.**
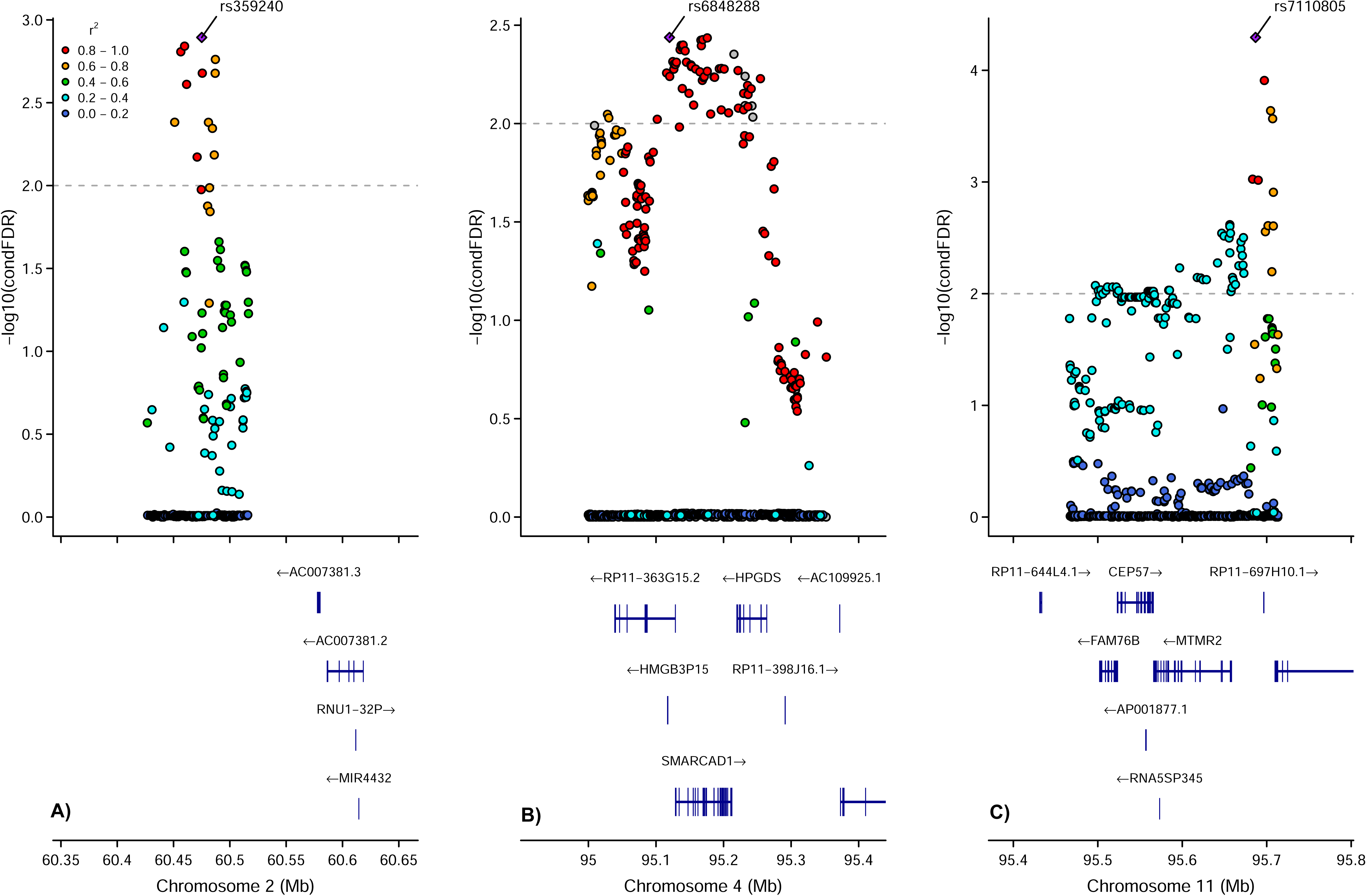
The genetic context of the strongest associations identified in the conditional false discovery rate (condFDR) analysis. Values for variants occupying the locus are shown on the left *y*-axis as –log10(condFDR). In each subplot, a single nucleotide polymorphism (SNP) with the strongest association is shown in the large purple square. The colour of the remaining markers reflects the degree of linkage disequilibrium (LD) with the strongest-associated SNP measured as r^2^ coefficient (described in the legend). The dotted line indicates the condFDR threshold of 0.01. A: surrounding of rs359240 (condFDR = 1.28 × 10^−3^). B: surrounding of rs6848288 (condFDR = 3.65 × 10^−3^). C: surrounding of rs7110805 (condFDR = 5.10 × 10^−5^).

Functional annotation of the candidate SNPs (see Supplementary Table 8) in the identified loci using FUMA revealed that most candidate SNPs were intronic and intergenic. We found 7 exonic candidate SNPs located on the chromosome 11 locus, and two of these SNPs were non-synonymous. Among the identified loci, 14 candidate SNPs had a CADD-score higher than 12.37 (a threshold suggested to reflect deleteriousness) (56). Full results of the FUMA analysis are presented in the Supplementary Table 9.

### Evaluation of the Identified Loci in an Independent Sample

We examined the identified loci in the association summary statistics from the independent TV-UKB and PC-UKB GWASs (11). We also evaluated the respective genetic correlations. PC-UKB was significantly correlated with gaming, but not with the other MoBa phenotypes, whereas TV-UKB showed positive genetic correlations with three screen behaviours in MoBa (r_g_ = 0.38–0.52, see Supplementary Table 13). Positive genetic correlations warrant evaluation of identified loci in TV-UKB and PC-UKB, despite considerable differences between the MoBa and the UKB cohorts (i.e. highly distinct age groups, and different in phenotypes *per se*).

Locus 3, represented by rs7110805, has the same direction of effect in the MoBa (social media use) and TV-UKB samples, with *p* < 0.05. Locus 1 and locus 2, represented by rs359240 and rs6848288, respectively, have the same direction of effect in the MoBa (social media use) and PC-UKB samples. Moreover, the *p*-value for rs359240 in the PC-UKB sample was nominally significant (*p* < 0.05). These positive evaluation results reassure validity of the identified loci.

### Genetic Overlap with Key Mental Traits and Disorders

We evaluated pairwise genome-wide genetic correlations between the three screen-based phenotypes with significant LDSC estimated SNP-heritability (TV watching, gaming, social media use) and six major psychiatric disorders, as well as EA. In addition, we estimated genetic correlations between the screen-based phenotypes themselves. The results are shown in Figure 4, and in Supplementary Tables 10-12. We corrected for multiple comparisons using FDR < 0.05.

**Figure 4.**
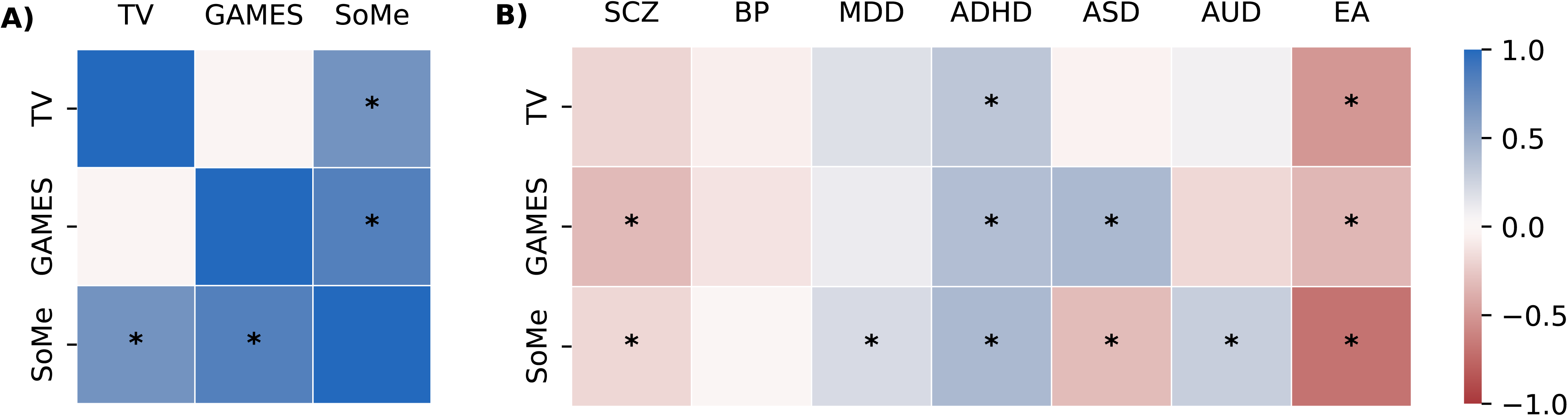
Genetic correlation estimates **A)** among screen behaviours and **B)** between screen behaviours and six major psychiatric disorders and educational attainment. Asterisks indicate significant estimates at FDR < 0.05 (Benjamini-Hochberg procedure). TV: watching movies/series/TV; GAMES: playing games on PC, TV, tablet, mobile, etc.; SoMe: communicating with friends on social media; SCZ, schizophrenia; BP, bipolar disorder; MDD, major depressive disorder; ASD, autism spectrum disorder; ADHD, attention-deficit hyperactivity disorder; AUD, alcohol use disorder; EA, educational attainment.

We identified significant genetic correlations between several screen time measures. Specifically, social media use was positively correlated with gaming (r_g_ = 0.83, SE = 0.27, *p* = 0.0021) and TV watching (r_g_ = 0.69, SE = 0.25, *p* = 0.0065), while TV and gaming were not significantly correlated.

We estimated significant genetic correlations between screen behaviours and psychiatric disorders (r_g_ in range 0.21–0.42). ADHD showed moderate positive genetic correlations with TV watching (r_g_ = 0.33, SE = 0.12, *p* = 0.006), gaming (r_g_ = 0.39, SE = 0.13, *p* = 0.0036), and social media use (r_g_ = 0.42, SE = 0.09, *p* = 3.67×10^−6^). ASD was positively correlated with gaming, but negatively with social media use. Both MDD and AUD were positively correlated with social media use (r_g_ = 0.21, SE = 0.065, *p =* 0.0012, and r_g_ = 0.31, SE = 0.12, *p =* 0.020, respectively), while SCZ was negatively correlated with gaming (r_g_ = −0.30, SE = 0.12, *p =* 0.0004). Finally, EA showed significant negative genetic correlation with all three screen behaviours, most strongly with social media use (r_g_ = −0.69, SE = 0.097, *p* = 9.38×10^−13^).

As a sensitivity analysis, we re-estimated all genetic correlations using GWAS on screen-based phenotypes based on subsample of participants without a history of any psychiatric disorder. The genetic correlation estimates in the two samples were highly concordant (Pearson correlation coefficient 0.98; see Supplementary Table 11, Supplementary Figure 4).

## Discussion

The present study investigated the genetic architecture of screen behaviours among adolescents and their associations with mental disorders and educational attainment (EA). Leveraging one of the largest birth cohorts in the world (33), we demonstrate that screen behaviours are heritable, highly polygenic traits. Furthermore, we identified the first genomic loci associated with adolescent social media use. We also demonstrate that television watching, gaming, and social media use share genetic signals with EA and major psychiatric disorders.

According to our results, three screen behaviors – television watching, gaming, and social media use – display significant nonzero *h^2^_SNP_* (Figure 1, Supplementary Table 3), which are generally in line with estimates for other behavioural traits (57–59). Furthermore, our estimates fall within the same range, but have more narrow standard errors than *h^2^_SNP_* reported for time spent on gaming, video watching and total screen time among children (8). To our knowledge, *h^2^_SNP_* estimates of social media use have not been reported before. Therefore, our results provide novel insights into the mechanisms behind screen behaviours.

Our study provides new perspectives on the shared genetic basis between screen time use and mental-health related phenotypes. We demonstrate that each of the screen behaviors displayed significant genetic correlations with one or more major psychiatric disorders (Figure 4). The most compelling pattern of associations was observed for ADHD, which has positive generic correlations with all three screen-based phenotypes. Although many studies have linked ADHD and problematic screen usage on a phenotypic level (60,61), genetic studies on the topic are scarce. Overall, our findings are consistent with previous results indicating that higher genetic liability for ADHD can contribute to longer screen time utilization, and that phenotypic association between screen time use and attention problems is partially explained by genetic factors (8,23,62,63). Notably, social media use displayed positive genetic correlation with both MDD and AUD, in addition to ADHD, but was negatively correlated with ASD. The mechanisms by which genetic factors may increase the susceptibility to both mental illness and screen behaviors remain to be uncovered.

Another distinct pattern was observed for EA, which displayed highly significant negative genetic correlations with television watching, gaming, and social media use. The relationship between decline in academic performance and increased screen time is well documented (17,64), but there are no studies investigating the potentially shared genetic background underlying this association. Our results suggest that adolescents with a high load of common genetic variants predisposing to problematic screen use may also be at higher risk for lower EA. While we cannot conclude on the causal relationship underlying these associations, or the specific mechanisms involved, we can hypothesize that the association between screen use and EA may at least partly be mediated by attention difficulties, which is consistent with the significant genetic correlations observed for both EA and screen behaviors with ADHD. Indeed, sustained attention is a key cognitive ability that is critical to successful goal-oriented behavior and is linked to academic achievement (65). Children genetically predisposed to attention difficulties are not only prone to more problematic screen use but may face an increased risk of lower educational outcomes (62,66,67). Further studies are necessary to investigate the possible link with attention problems. Our sensitivity analysis indicate that the identified associations were not driven by participants with a history of mental illness in the screen behaviour GWASs (Supplementary Figure 4). Based on the current findings, we suggest that individuals with a high load of genetic risk factors for a particular psychiatric disorder (but not necessarily with the diagnosis itself) may be at higher risk for displaying more extreme screen behaviours.

By combining GWAS summary statistics on social media use and EA (32) in the condFDR analysis (34,35), we enhanced discovery in the moderately powered social media use GWAS, and identified three genomic loci associated with social media use (Figure 2, Supplementary Table 8). The most significant variant (rs7110805, intergenic) is located on chromosome 11 and represents a broad region of associations (Figure 3C), which contains three protein-coding genes: *MTMR2, FAM67B,* and *CEP57. MTMR2* encodes myotubularin-related protein 2 and is moderately expressed in the brain(68), mostly in oligodendrocytes and neurons. Its neural functions remain unclear, though synaptically localized *MTMR2* might maintain excitatory synapses by inhibiting excessive endosomal production and destructive trafficking to lysosomes (69). According to the Open Targets Genetics model (43,54), *MTMR2* is also predicted to be likely casual (L2G pipeline scores > 0.6) for EA (70,71), consistent with our hypothesis about shared genetic factors between social media use and EA. *FAM67B* gene encodes its eponymous protein, with low brain expression levels. Nevertheless, it has a moderate chance to be casual at locus discovered in the GWAS of vertex-wise sulcal depth (L2G score = 0.54) (72). Finally, the *CEP57* gene, which encodes centrosomal protein 57, is moderately expressed in neurons and fetal astrocytes (68). The Cep57 protein controls centriole duplication and centrosome maturation for faithful cell division. Interestingly, *CEP57* has more than a 50% chance to be casual at loci discovered in GWAS for leisure television watching, cortical surface area, and cognitive aspects of EA (11,73,74). Again, these findings are consistent with our hypothesis of shared genetic basis between EA and screen behaviors.

The second locus associated with social media use is represented by an intergenic variant rs359240 on chromosome 2, with no protein-coding genes in the direct vicinity (Figure 3A). Nevertheless, this variant has a CADD score of 19.6, suggesting high deleteriousness(56). Interestingly, rs359240 crossed genome-wide suggestive significance threshold for several brain-related and behavioral traits, like smoking, risk-taking behavior, and age at first sexual intercourse and age at first live birth(59,75,76).

Finally, the third identified locus was located on chromosome 4, and is represented by an ncRNC intronic variant rs6848288, which has multiple LD-linked variants with low condFDR values (Figure 3B). The OpenTargets platform links rs6848288 most strongly to the *SMARCAD1* gene*. SMARCAD1* encodes its eponymous protein and participates in transcriptional regulation, heterochromatin maintenance, DNA repair, and replication, though the molecular basis of its role in these processes is not fully understood (77). The *SMARCAD1* gene is weakly expressed in both human astrocytes and neurons (68). However, according to the L2G pipeline, this gene has more than a 70% chance to be causal at loci discovered in GWAS for cortical thickness and vertex-wise surface area (72,74). The link between putative causal genes for screen behaviors and brain structures measures is intriguing, especially in light of studies that identified associations between internet gaming disorder and structural and functional brain changes (78). Brain changes associated to a more moderate level of digital media use remain to be investigated, and further studies in adolescent population are warranted.

Functional characterization of the identified loci by FUMA revealed two exonic nonsynymous SNPs (i.e., with a high probability of deleteriousness, as these variants change the produced protein’s amino acid sequence) located on chromosome 11 (Supplementary Table 9). However, like in GWAS on other complex human traits phenotypes, most of the identified candidate SNPs reside in non-coding DNA, suggesting a regulatory and more indirect influence on the phenotype (79). This may also suggest that the loci do not influence a distinct biological process but represent non-specific genetic effects common to several mental-health related phenotypes. These findings may also reflect insufficient statistical power. Nevertheless, follow-up studies are warranted to determine the specific causal genetic variants underlying the loci detected here.

Replication of the identified variants for social media use was not possible due to the absence of comparable independent datasets. Nevertheless, we examined the identified loci in the association summary statistics from the TV-UKB and PC-UKB GWAS conducted in the UKB cohort. The lead SNPs rs359240 and rs7110805 had the same direction of effect as in the MoBa cohort and nominally significant *p*-values in the PC-UKB and TV-UKB samples, respectively. These findings are notable, given the considerable heterogeneity between the cohorts and different phenotypes. Nevertheless, we acknowledge the possibility that these results may reflect random variation rather than a definitive pattern and recognize this as a limitation.

Our study is not without limitations. Generally, selection bias is a major challenge in cohort studies, and MoBa participants were found to not be representative of the entire Norwegian population (80). Moreover, we were not able to estimate potential discrepancies between self-reported and objectively measured screen time, and we did not have information about the content participants were engaging with during their screen use. The complex associations between phenotypes (attention difficulties and EA, as well as socioeconomic factors that might be linked both to EA and screen use, etc.) prevents us from translating the observed genetic correlations into actual pleiotropy. The initial set of single-trait GWASs performed in our study did not unambiguously identify any loci associated with screen behaviours, though many variants reached the suggestive threshold. We hold the view that this pattern of results is merely due to low power, despite the sample being substantially larger than any prior study of screen behaviours. Insufficient statistical power prevents us from using tools that can be highly valuable in identifying causal relationships or studying genetic overlap between traits (e.g., Mendelian Randomization and MiXeR). Therefore, we urge the research community to continue collecting large-scale data about screen-based activities, as it will greatly improve our understanding of one of the most widespread behavioural traits in the modern society.

In summary, we demonstrated that television watching, gaming, and social media use are heritable, highly polygenic traits, which display significant genetic correlations with one or more major psychiatric disorders, and are negatively correlated with EA. Furthermore, we identified three genomic loci associated with adolescent social media use. Overall, our results provide new insights into the genetics of screen behaviours and may generate new hypotheses regarding the relationship between screen time use, mental health, and EA during adolescence.

## Data Availability

The datasets supporting the conclusions of this article are available from the Norwegian Institute of Public Health, but restrictions apply to the availability of these data. The study website provides details on how to access data and information on the available variables (https://www.fhi.no/en/ch/studies/moba/for-forskere-artikler/research-and-data-access/).

## Acknowledgements

The Norwegian Mother, Father and Child Cohort Study is supported by the Norwegian Ministry of Health and Care Services and the Ministry of Education and Research. We are grateful to all the participating families in Norway who take part in this on-going cohort study. We also thank deCODE genetics for genotyping of the main part of the MoBa sample.

This work was performed on Services for sensitive data (TSD), University of Oslo, Norway, with resources provided by UNINETT Sigma2 - the National Infrastructure for High Performance Computing and Data Storage in Norway.

