## Supplementary Figures for "Genome-wide analysis of screen behaviors among adolescents identifies novel loci and overlap with educational attainment and mental disorders"

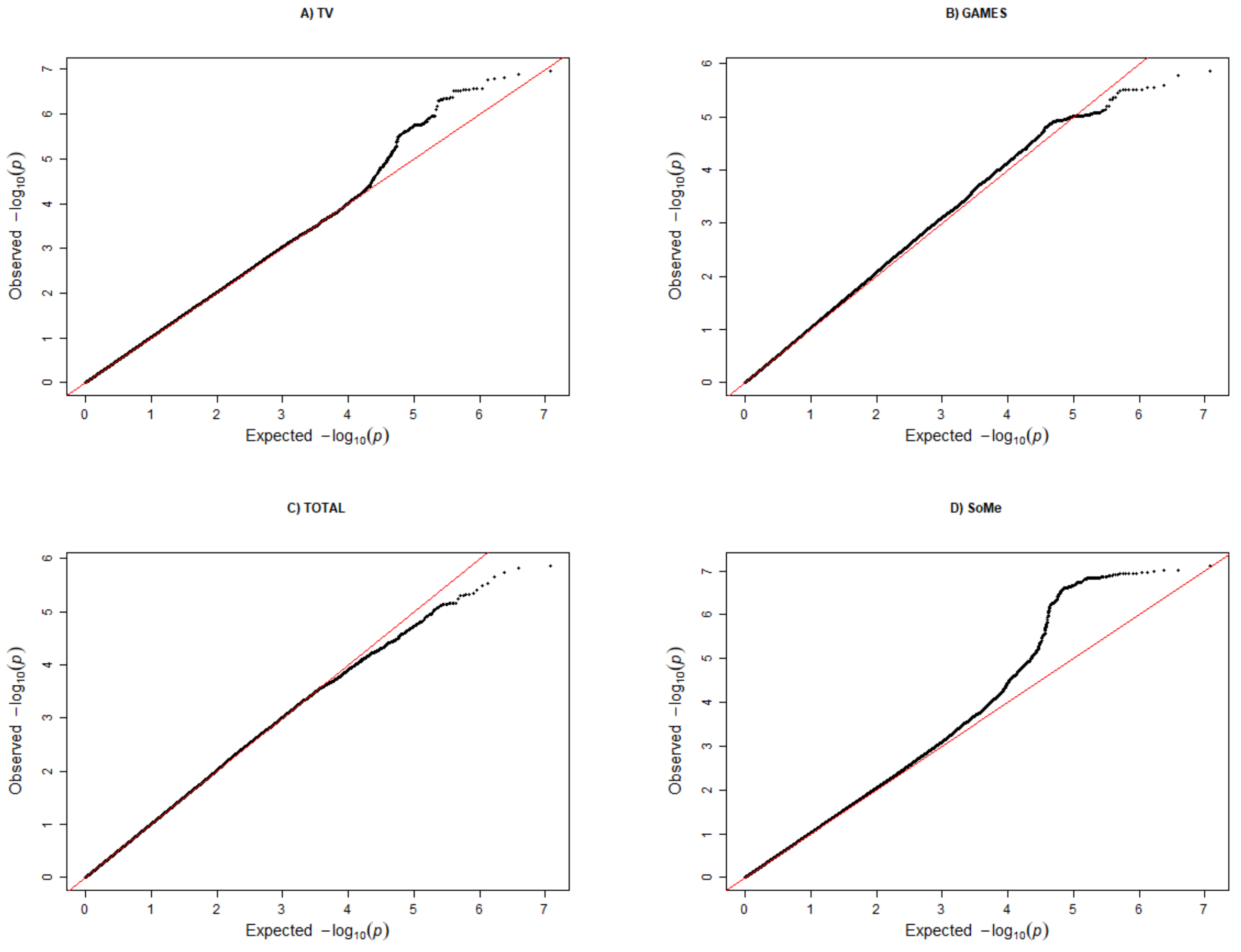


**Supplementary Figure 1.** Quantile-Quantile plots of *p*-values from genome-wide association studies of the screen behaviors in the MoBa cohort (*n* = 16 027).

TV: watching movies/series/TV; GAMES: playing games on PC, TV, tablet, mobile, etc.; TOTAL: sitting/lying with PC, mobile, or tablet; SoMe: communicating with friends on social media.

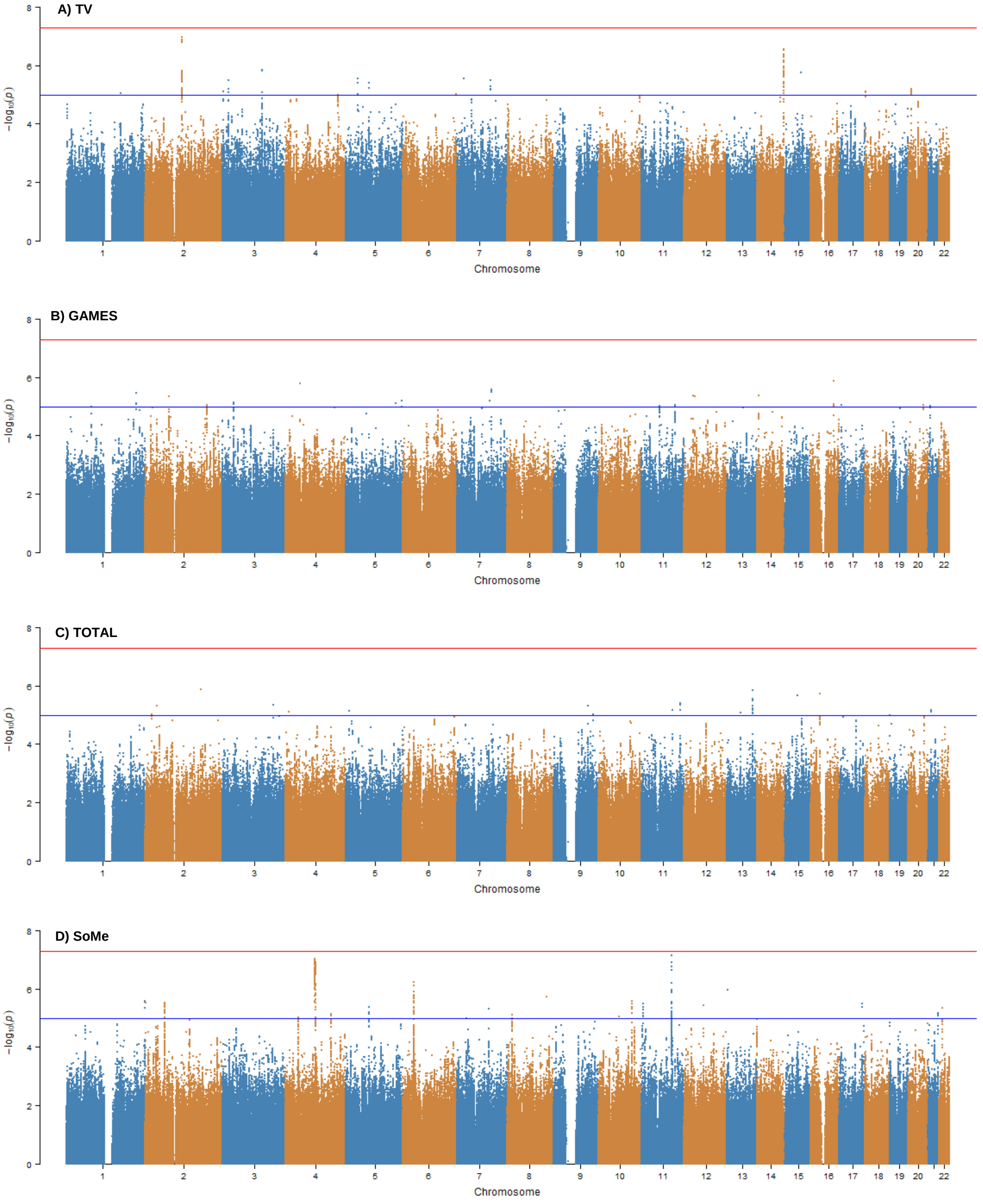


**Supplementary Figure 2.** Manhattan plots of genome-wide association studies of the screen behaviors in the MoBa cohort ($n$ = 16,027).

The $x$-axis shows genomic position (chromosomes 1–22), and the $y$-axis shows statistical significance as –log10(p-value). The blue line indicates the genome-wide suggestive threshold ($p < 1 \times 10{-}5$), the red line indicates the genome-wide significance threshold ($p < 5 \times 10{-}8$).

TV: watching movies/series/TV; GAMES: playing games on PC, TV, tablet, mobile, etc.; TOTAL: sitting/lying with PC, mobile, or tablet; SoMe: communicating with friends on social media.

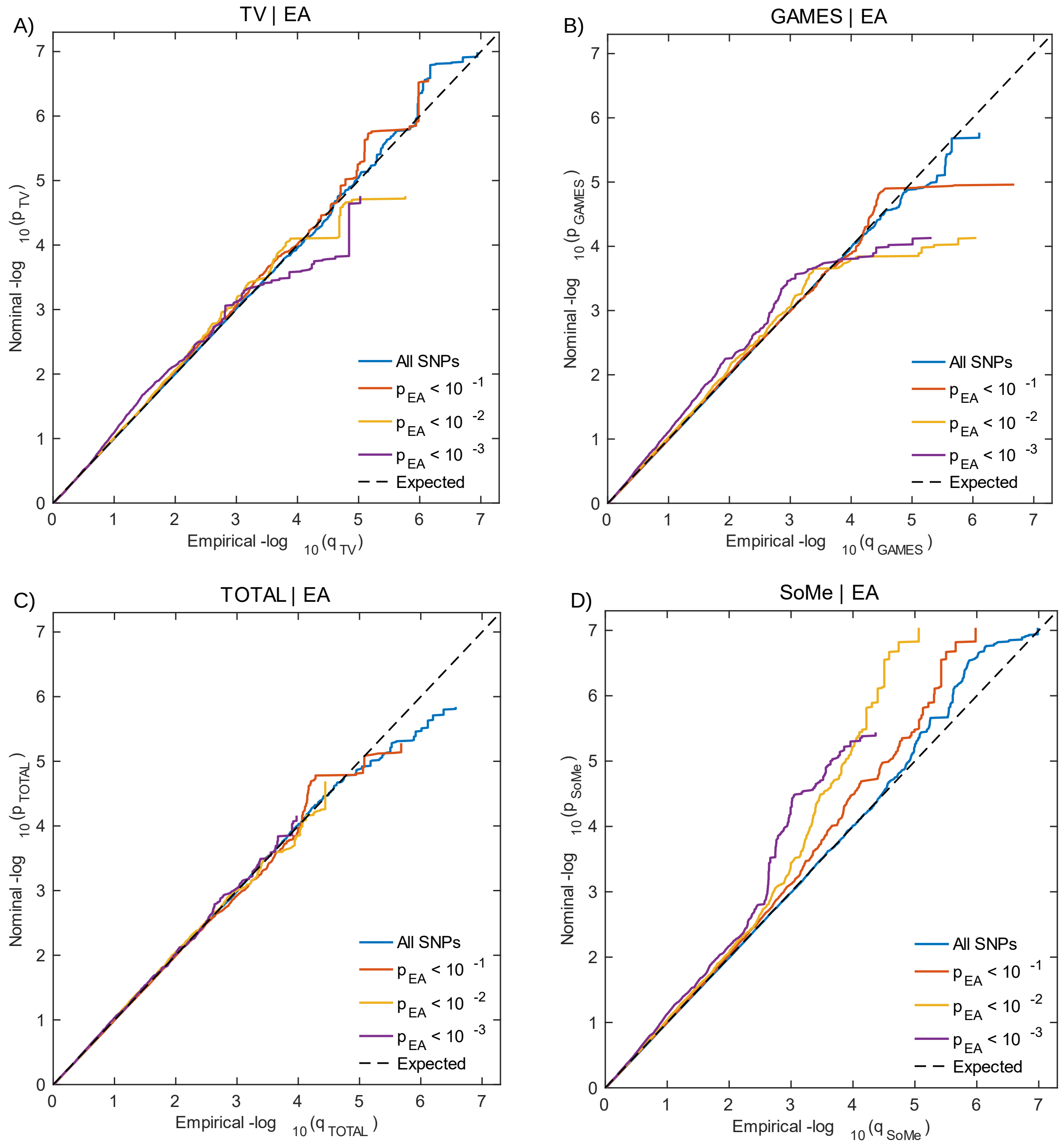


**Supplementary Figure 3.** Conditional Quantile-Quantile (QQ) plots.

QQ plots demonstrate relation between expected (*x*-axis) and observed (*y*-axis) significance of markers in the primary trait when markers are stratified by their *p*-values in the conditional trait.

Screen time use phenotypes from the MoBa cohort are conditioned on the educational attainment (EA) phenotype. A sequence of four nested strata is presented: all single nucleotide polymorphisms (SNPs) (i.e. *p*-values of the secondary trait ≤ 1.00) (blue), $p_{\text{conditional\_trait}} < 0.1$ (red), $p_{\text{conditional\_trait}} < 0.01$ (yellow), and $p_{\text{conditional\_trait}} < 0.001$ (purple). Dashed black line demonstrates expected behaviour under no association. Increasing degree of leftward deflection from the no-association line for strata of SNPs with higher significance in the conditional trait indicates polygenic overlap.

TV: watching movies/series/TV; GAMES: playing games on PC, TV, tablet, mobile, etc.; TOTAL: sitting/lying with PC, mobile, or tablet; SoMe: communicating with friends on social media.

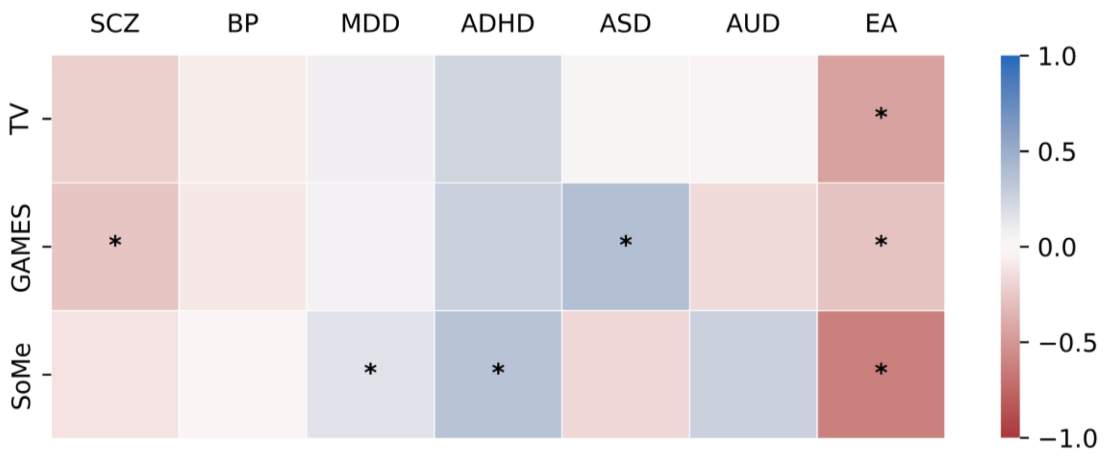


**Supplementary Figure 4.** Genetic correlation estimates between screen-based behaviors and six major psychiatric disorders and educational attainment, subsample of participants without a history of any psychiatric disorder (n = 13 375).

Asterisks indicate significant estimates at FDR < 0.05 (Benjamini-Hochberg procedure).

TV: watching movies/series/TV; GAMES: playing games on PC, TV, tablet, mobile, etc.; SoMe: communicating with friends on social media; SCZ, schizophrenia; BP, bipolar disorder; MDD, major depressive disorder; ASD, autism spectrum disorder; ADHD, attention deficit hyperactivity disorder; AUD, alcohol use disorder; EA, educational attainment.
